# Biomarker Heterogeneity in Patients Receiving Lecanemab in the Real World

**DOI:** 10.64898/2025.12.10.25341811

**Authors:** Masanori Kurihara, Ryoko Ihara, Gen Yoshii, Ryosuke Shimasaki, Keiko Hatano, Taro Bannai, Fumio Suzuki, Kenji Ishibashi, Ko Furuta, Katsuya Satoh, Aya Midori Tokumaru, Kenji Ishii, Atsushi Iwata

## Abstract

**Background:** While amyloid-β (Aβ) biomarker positivity is sufficient before initiating anti-Aβ antibody therapy, recent revised criteria also highlight the importance of other biomarkers (ATNIVS) to understand heterogeneity in AD.

**Design, Setting, and Participants:** We reviewed patients who attended our specialty clinic between December 2023 and October 2024. Some participated in tau PET study (^18^F-MK6240). MRI was assessed using Fazekas score. Remaining samples were analyzed for plasma neurofilament light chain (NfL), glial fibrillary acidic protein (GFAP), and CSF α-synuclein seed amplification assay (SAA).

**Results:** During the period, 200 attended and 147 proceeded to screening. Lecanemab was started in 93 of 108 A+ patients; mean age 74.2 years, 73.1% female. While all tested started on lecanemab were positive on amyloid PET, 21% had only regional positivity with lower Aβ burden (centiloid 31.3 ±17.5 vs 67.6 ±20.2) and higher age (79.2 ±5.1 vs 73.3 ±8.9). While all tested had CSF Aβ42/40 values below the single cut-off 0.067 in Japan, three (8.6%) had values close to the cutoff (0.059–0.067), all of whom were all male. Other biomarkers also widely varied from normal to fully abnormal; CSF pTau181 (40.5–168 pg/mL, cut-off 56.5), tau PET-based Braak stage (0–VI), NfL (10.0–103.3 pg/mL), GFAP (121.9–652.5 pg/mL), Fazekas score (0–3), and positive α-synuclein SAA (25–33%). Some associations were indicated including higher Fazekas scores in amyloid PET regional-positive group and higher plasma NfL in CSF Aβ42/40 0.059–0.067 group.

**Conclusions:** We identified substantial heterogeneity in ATNIVS biomarker profiles among patients receiving lecanemab in a real-world setting.

## 1 Introduction

Alzheimer’s disease (AD) is a neurodegenerative disorder characterized by senile plaques and neurofibrillary tangles (NFT) in the brain, which are aggregates of amyloid β (Aβ) and tau, respectively. As Aβ aggregation is considered the main upstream factor of AD pathophysiology [1], disease-modifying therapy (DMT) targeting Aβ has been developed, and anti-Aβ antibodies lecanemab and donanemab finally proved clinical efficacy in phase 3 trials [2, 3].

After the publication of the phase 3 trial results [2] and approval by the Food and Drug Administration in the United States (US-FDA), Japan was the second country to approve lecanemab on September 25, 2023, by the Ministry of Health, Labor and Welfare. After national health insurance coverage and release of the national optimal use guidelines (OUGs) [4, 5], lecanemab became commercially available on December 20, 2023. We immediately initiated lecanemab infusion on December 25, 2023. Although most hospitals required additional preparation time, the number of centers prescribing lecanemab has steadily increased in Japan. Lecanemab has been approved by many other countries and unions, including China, Korea, Israel, the United Arab Emirates, the United Kingdom, and the European Union.

Although confirmation of Aβ pathology is sufficient to initiate anti-Aβ antibody therapy in clinical settings, recently revised criteria highlight the importance of evaluating biological heterogeneity, including tau and co-pathologies, in research settings [6]. In line with the eligibility for anti-Aβ antibodies, recently revised criteria for the diagnosis of AD require abnormality in validated core 1 biomarkers, including amyloid positron emission tomography (PET) and cerebrospinal fluid (CSF) Aβ42/Aβ40 [6]. Clinically, clinical assessment and positivity of core 1 biomarkers are considered sufficient for diagnosing symptomatic AD. However, Aβ biomarker abnormality (≈ Aβ pathology in the brain) increases with age and thus becomes prevalent in cognitively normal or mild cognitive impairment(MCI)/dementia owing to other causes, particularly in the older population [7, 8]. Therefore, Aβ biomarker abnormality does not necessarily mean that a patient’s cognitive impairment is mainly caused by AD, and therefore, clinical judgement is always important [9].

The revised criteria also propose updated biological staging using tau PET, which would be useful for understanding the association between biological and clinical staging [6]. Although no tau PET tracer is currently covered by health insurance in Japan, continued research is important to understand tau PET distribution and the concordance/discordance of biological/clinical AD stages. The revised criteria also updated biomarker categories important in understanding the AD pathophysiological process known as ATNIVS (A, amyloid; T1, phospho/secreted tau; T2, AD tau proteinopathy; N, neurodegeneration; I, inflammation; V, vascular; and S, α-synuclein (α-syn)). Therefore, although not required for the clinical diagnosis of AD or for eligibility assessments of DMTs, measurement of these biomarkers in real-world patients receiving lecanemab is important to understand the biological heterogeneity within patients that could affect disease progression, adverse effects, and clinical efficacy.

Therefore, this study aimed to evaluate the heterogeneity of the ATNIVS biomarkers in patients receiving lecanemab therapy in a real-world setting.

## 2 Methods

### 2.1 Study Population and Design

We reviewed all patients who visited our AD DMT clinic between December 2023 and October 2024. We selected this period in this study, which was before the modification of our initial screening requirement in November 2024, to include the Mini-Mental State Examination (MMSE) 20–21 owing to the launch of donanemab on November 26, 2024. The requirement for referral included a diagnosis of MCI or mild dementia, MMSE ≥22 (mandatory by Japanese OUGs), no evidence of disease other than AD primarily causing cognitive impairment, and the ability to undergo magnetic resonance imaging (MRI) scans. Those interested in treatment subsequently underwent 3T MRI, clinical dementia rating (CDR; also mandatory by the Japanese OUG), and amyloid PET or CSF biomarker testing.

### 2.2 Ethical Approval and Consent

This study was performed in accordance with the tenets of the Declaration of Helsinki and approved by the Institutional Review Board of the Tokyo Metropolitan Institute for Geriatrics and Gerontology (TMIG) (R23-117). Tokyo Medical Biobank (R21-038), α-syn real-time quaking-induced conversion (RT-QuIC) (R24-032), and ^18^F-MK6240 PET studies (R22-088) were also approved. Written informed consent was obtained for CSF biomarker measurements, ApoE phenotype testing, and biobank and PET studies.

### 2.3 MRI Protocol and Fazekas score

All MRI scans were obtained using the same 3T MRI scanner (Ingenia Elition 3.0T; Koninklijke Philips NV, Amsterdam, Netherlands). Diffusion-weighted imaging, susceptibility-weighted imaging, fluid-attenuated inversion recovery, and T2-weighted and 3D T1-weighted imaging were performed in accordance with the Japanese guidelines [10]. The presence or absence of contraindication, including microhemorrhages ≥5, macrohemorrhage, superficial siderosis, or vasogenic edema/sulcal effusion, was evaluated by radiologists specialized in neuroradiology, and was also confirmed in a consensus conference. White matter hyperintensities on FLAIR images were scored in the periventricular (PV) and deep white matter by an expert (A. M. T.) using the Fazekas score [11]. Although not recommended in the appropriate use recommendations from US experts [12], a Fazekas score of 3 was not an exclusion criterion in Japanese OUGs; thus, lecanemab was initiated after informing the risk if deemed necessary.

### 2.4 Amyloid PET scan and Quantification

During this period, all amyloid PET scans were performed using ^18^F-flutemetamol (Vizamyl; Nihon Medi-Physics, Tokyo, Japan). Static emission data were acquired 90–110 min after IV bolus injections. The administered dose was approximately 185 MBq. Visual reads were obtained by an expert (K. Ishii). According to the approved method, PET images were visually interpreted in five brain regions (parietal, frontal, temporal lobes, striatum, and posterior cingulate cortex/precuneus), and the result was considered positive if there was uptake in any region. Centiloid scales were calculated using a previously reported automated semi-quantitative analysis technique without anatomical images [13] provided by the VIZCalc software (Nihon Medi-Physics). The results of those using previous amyloid PET via approved but different tracers or scanning protocols were reconfirmed to be visually positive by our expert reading, but were not included in the quantification analyses.

### 2.5 CSF AD Biomarker Measurement

CSF was obtained via a standard lumbar puncture. During the study period in Japan, the Aβ42/Aβ40 ratio measured using the LUMIPULSE system (FUJIREBIO INC., Tokyo, Japan) and its assays were the only approved CSF assay to confirm eligibility for lecanemab. The CSF concentrations of Aβ42, Aβ40, phosphorylated tau 181 (pTau181), and total tau (tTau) were measured using the LUMIPULSE system and its assays [14, 15] at SRL, Inc. (Tokyo, Japan). Aβ42 and Aβ40 measurements were covered by health insurance, whereas pTau181 and tTau were research-funded. Predetermined cutoffs were Aβ42/Aβ40 0.067 [15], pTau181 56.5 pg/mL, and tTau 404 pg/mL [14]. Although Japan uses a single cutoff of Aβ42/Aβ40 0.067, the US FDA suggested two cutoff values, and the intermediate range 0.059– 0.072 was considered a likely-positive range that may be associated with some uncertainties [16].

We previously confirmed that CSF pTau181 mildly but significantly increases with Aβ pathology alone [17]. However, pTau181 was significantly higher in those with Aβ plus tau pathology (≥ Braak neurofibrillary tangle stage III), and a predictive cutoff for Aβ plus tau pathology was 83.1 pg/mL, based on our previous autopsy data and conversion formula between Innotest ELISA and Lumipulse assay [14, 17]. Those who consented were tested for the ApoE phenotype by isoelectric focusing followed by western blotting [18]. *APOE* testing was not covered by insurance or recommended in Japan, and the remaining patients were not tested.

### 2.6 TMIG Biobank

Patients were recruited to our biobank during their first visit to the Memory Clinic, before lumbar puncture, or before lecanemab treatment. For patients who underwent lumbar puncture, the remaining CSF samples were handled and stored as previously described [14, 19]. Blood collection was also conducted, and plasma samples were collected in 2Na-EDTA tubes, centrifuged at 2,400 × *g* for 15 min within 30 min, and stored at −80 °C for future research.

### 2.7 α-syn Seed Amplification Assay (SAA)

Frozen CSF samples in the biobank available on July 19, 2024, were used for α-syn SAA analyses via RT-QuIC using previously published methods [19]. The tester was blinded to any clinical or biomarker information, and positivity was determined based on predetermined criteria [19].

### 2.8 Plasma NfL and GFAP measurement

The remaining plasma samples in the biobank were analyzed for neurofilament light chain (NfL) and glial fibrillary acidic protein (GFAP). Measurements were conducted using the Simoa HD-X platform and the Neurology 2-Plex B multiplex assay (Quanterix, Billerica, MA, USA) at Raybiotech, Inc. (Norcross, GA, USA) after a 4-fold dilution. The measurements were conducted in duplicate, and the average concentration was used.

### 2.9 ^18^F-MK6240 PET and Tau PET-based Braak Staging

Emission data were acquired 90–120 min after intravenous ^18^F-MK6240 administration as previously reported [20]. PET images were visually evaluated by two experts (K. Ishibashi and K. Ishii), and PET-based Braak staging was conducted in reference to published methods [21–23] as previously reported [24, 25].

### 2.10 Statistical methods

Statistical analyses were conducted using GraphPad Prism version 9 (GraphPad Software, San Diego, CA, USA) or R version 4.2.1 (R Foundation for Statistical Computing, Vienna, Austria) and the graphical interface EZR (Saitama Medical Center, Jichi Medical University, Saitama, Japan) [26]. Missing data were addressed using a pairwise deletion approach. Categorical variables are expressed as percentages, and differences between groups were evaluated using Fisher’s exact test. Normally distributed continuous variables are expressed as mean ± standard deviation, and differences between groups were tested using the Student’s *t*-test or one-way analysis of variance (ANOVA), followed by *post hoc* analyses using Tukey’s test. Continuous variables without a normal distribution were expressed as median (interquartile range), and differences between groups were tested using the Mann–Whitney *U* test. Correlations were evaluated using Pearson’s method. Receiver Operating Characteristic (ROC) analysis was conducted for the centiloid scale, and cutoffs were determined as the values that maximized the Youden’s index.

## 3 Results

### 3.1 Baseline characteristics of the patients receiving lecanemab

During this period, 200 patients visited our DMT clinic, and 147 underwent further screening. Fourteen had preexisting positive amyloid biomarkers (A+), and seven failed screening before biomarker testing (contraindication on MRI, CDR 0). Of the remaining 126 patients, 80 (63%) underwent amyloid PET, and the remaining selected cerebrospinal fluid (CSF). A+ was confirmed in 74% of the cases by PET and 76% by CSF. Thirty-two were non-eligible because of a negative amyloid biomarker (A−) and included as controls. Of the 108 A+ patients, lecanemab was initiated in 93. Baseline characteristics compared with those in previous reports [2, 27–32] are summarized in **Table 1**. The mean age was slightly higher than that in the phase 3 trial (71.4), consistent with real-world US data [28, 30, 31].

**Table 1.** Baseline characteristics compared to the phase 3 study and other recent reports of real-world data. Values higher than those of the Phase 3 study are highlighted in orange, and values lower are highlighted in blue. Compared to the phase 3 study and other real-world data, the characteristics of the patient population in this study were a high female proportion and, low ApoE ε4 carrier rate. Although not reported in other real-world data, the amyloid PET centiloid level was lower than that reported in the phase 3 trial.

|  | Phase 3 Clarity AD |  | TMIG | WashU<br>USA (n=234)<br>Paczynski<br>JAMA Neurol<br>2025 | NNI<br>USA (n=71)<br>Shields JPAD<br>2024 | Komodo Data<br>USA (n=3155)<br>Sabbagh<br>Neurol Ther<br>2025 | Veteran<br>USA (n=56)<br>Mittler<br>J Neurol<br>2025 | TLVMC<br>Israel (n=86)<br>Bregman<br>Als Res Ther<br>2025 |
| --- | --- | --- | --- | --- | --- | --- | --- | --- |
|  | Global<br>(n = 859)<br>van Dyck<br>NEJM 2023 | Asia<br>(n = 146)<br>Chen et al.<br>JPAD 2025 | This study<br>(1y, n = 93) |  |  |  |  |  |
| Age | 71.4 ±7.9 | 70.3 ±8.1 | 74.2 ±8.1 | 74.4±6.7 | 72 | 75.0±6.8 | 75.9±5.3 | 72.0±8.2 |
| Sex (female) | 51.6% | 58.2% | 73.1% | 50% | 62% | 55.8% | <10% | 53% |
| MCI | 61.5% | 69.9% | 62.4% |  | 49.3% | 60.8% | 48.2% | 72% |
| Global CDR 0.5 | 80.8% | 96.6% | 88.2% | 85% |  |  |  |  |
| CDR-SB | 3.17 ±1.34 | 2.76 ±1.00 | 2.99 ±1.31 |  |  |  |  |  |
| MMSE | 25.5 ±2.2 | 25.2 ±2.1 | 24.7 ±2.0 | 24±4 | 25 |  |  | 24.0±2.7 |
| Current use of<br>symptomatic | 52.0% |  | 41.9% |  | 94% | 67.6% | 48.2% |  |
| APOE ε4 carrier | 68.9% | 72.6% | 32.4%<br>(12/37) | 62.5% | 64% |  |  | 53% |
| PET centiloid<br>mean | 77.9 ±44.8 | 82.2 ±33.6 | 59.8 ±24.4 |  |  |  |  |  |
| range | -16.6 to 213 |  | 3.1 to 112 |  |  |  |  |  |

Although men comprised nearly half of the participants in the phase 3 trials [2], and real-world data from some institutions showed a similar tendency [28, 32], the proportion of women was as high as 73.1%, closer to the general AD population (men: women approximately 1:2) [33–35]. Only 32.4% were *APOE* ε4 carriers, which was lower than that of the phase 3 trials [2, 27] or reports from institutions in other countries [29, 32, 36]. Considering previous reports on the Japanese population [37], the low prevalence could be because of the small sample size for *APOE* status. Other baseline characteristics were similar to those observed in the phase 3 trial.

### 3.2 Amyloid (A) and Tau (T1, T2) biomarkers

Fifty-three patients underwent amyloid PET with flutemetamol at the time of evaluation. Although all patients who started lecanemab were positive for amyloid PET, 21% (11/53) had only regional positivity (A+_region_; 1–4 regions) compared to the widespread positive results (A+_wide_; all 5 regions) commonly observed in symptomatic AD. Among the regional group, Aβ deposition was most frequently observed in the frontal lobe, followed by the temporal lobe and posterior cingulate cortex/precuneus (**Figure 1A**). The mean age was significantly higher in the A+_region_ group compared to the A+_wide_ group (79.2 ±5.1 vs 73.3 ±8.9 years, p = 0.04). Other baseline characteristics did not differ significantly between the groups (**Table 2**). Overall, the amyloid burden measured using the centiloid scale (0 = young normal, 100 = typical AD dementia) ranged from 3.1 to 112. Centiloid values significantly increased from A−, A+_region_, to A+_wide_ (**Figure 1B**). ROC analysis identified optimal cutoff values of 10.9–20.8 for distinguishing A− vs A+ and 41.5 for A+_region_ vs A+_wide_ (**Figure 1C**).

**Figure 1.**
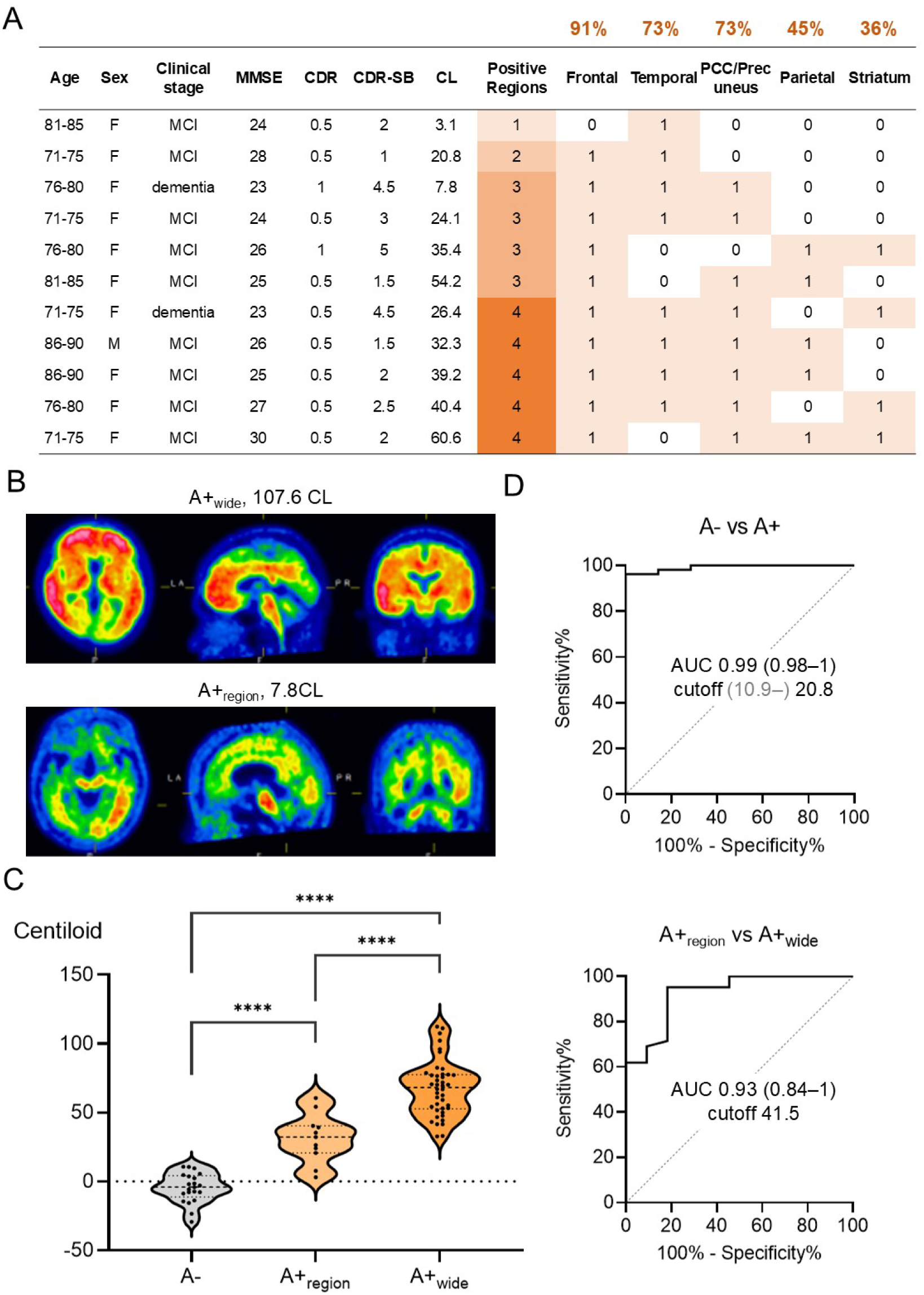
Regional amyloid PET positivity and centiloid scales of amyloid-positive patients on lecanemab (n = 53) and amyloid-negative patients (n = 21) A) Characteristics of 11 patients with regional (1–4 of 5) amyloid positivity. All patients were 72 or older in age, and amyloid deposition was most frequently observed in the frontal lobe, followed by the temporal lobe and posterior cingulate cortex/precuneus. B) Representative amyloid PET images of A+_wide_ and A+_region_. While Aβ deposition is observed in all five regions in A+_wide_, deposition was observed only in three regions (frontal, temporal, and PCC/precuneus in this case) in A+_region_. C) Centiloid scale significantly increased from A− (−4.6 ±10.7), A+_region (_31.3 ±17.5_)_, to A+_wide_ (67.6 ±20.2). D) ROC curves differentiating A− vs A+ or A+_region_ vs A+_wide_. The Centiloid scale had a high AUC for both differentiations. A−, amyloid negative; A+_region_, amyloid regionally positive; A+_wide_, amyloid widespread positive; AUC, area under the curve; CDR, clinical dementia rating; CDR-SB, clinical dementia rating sum of boxex; CL, centiloid scale; MMSE mini-mental state examination; PCC, posterior cingulate cortex

**Table 2.** Comparison between amyloid PET regional vs widespread positive and CSF Aβ42/40 likely vs clearly positive groups. A+_region_, amyloid regionally positive; A+_wide_, amyloid widespread positive; Aβ, amyloid-beta; CDR, clinical dementia rating; CDR-SB, CDR sum of boxes; CSF, cerebrospinal fluid; DWM, deep white matter; GFAP, glial fibrillary acidic protein; MCI, mild cognitive impairment; MMSE, mini-mental state examination; NfL, neurofilament light chain; pTau181, tau phosphorylated at threonine 181; PV, periventricular; SAA, seed amplification assay; αsyn, alpha-synuclein

| | | Amyloid PET (n = 53) | | | CSF Aβ42/40 (n =35) | | |
| --- | --- | --- | --- | --- | --- | --- | --- |
|  |  | A+region<br>(n = 11) | A+wide<br>(n = 42) | p value | 0.059–0.067<br>(n = 3) | < 0.059<br>(n = 32) | p value |
| Baseline | Age | 79.2 ± 5.1 | 73.3 ± 8.9 | * 0.040 | 76.7 ± 7.4 | 73.9 ± 7.5 | 0.55 |
|  | Female | 90.9% | 78.6% | 0.67 | 0% | 68.8% | * 0.044 |
|  | MCI | 81.8% | 54.8% | 0.34 | 66.7% | 59.4% | 1 |
| | MMSE | 25.5 ± 2.2 | 24.5 ± 2.1 | 0.17 | 23.3 ± 0.58 | 24.7 ± 1.9 | 0.26 |
|  | CDR 0.5 | 81.8% | 81.0% | 1 | 100% | 96.9% | 1 |
| | CSR-SB | 2.68 ± 1.38 | 3.32 ± 1.19 | 0.13 | 2.17 ± 0.76 | 2.86 ± 1.42 | 0.42 |
| T1 | CSF pTau181 | n/a | n/a | | 67.1 ± 24.5 | 98.0 ± 36.5 | 0.16 |
| T2 | tau PET-based Braak stage | 1 [0.5-1.5] | 4 [3.5-4.5] | 0.20 | 1 | 5 [5-6] | 0.21 |
|  | available data | 2/11 | 3/42 |  | 1/3 | 5/32 |  |
| N | plasma NfL | 23.4 ± 7.3 | 29.2 ± 20.7 | 0.40 | 30.8 ± 2.1 | 20.8 ± 6.6 | * 0.015 |
| I | plasma GFAP | 290 ± 148 | 348 ± 125 | 0.24 | 264 ± 139 | 312 ± 123 | 0.54 |
|  | available data | 10/11 | 29/42 |  | 3/3 | 28/32 |  |
| V | Fazekas DWM | 1 [1-2] | 1 [1-1] | † 0.067 | 2 [1.5-2] | 1 [1-1] | 0.13 |
|  | Fazekas PV | 2 [1-2.5] | 1 [1-1] | * 0.022 | 2 [1.5-2.5] | 1 [1-2] | 0.19 |
| S | CSF αsyn SAA | n/a | n/a | | 0% | 31.8% | 1 |
|  | available data |  |  |  | 1/3 | 22/32 |  |

Although CSF Aβ42/40 was below the cutoff of 0.067 in all patients who started on lecanemab, the value ranged from 0.027 to 0.063, including three with values close to the cutoff in the likely-positive range by the US FDA [16]. All three patients with CSF Aβ42/40 in this range were men. Other baseline characteristics did not differ significantly between groups (**Table 2**).

CSF pTau181 concentration ranged from 40.5 to 168 pg/mL (**Figure 2A**). Using the previously suggested cutoff of 56.5 [14] or the normal reference range of 21.5–59.0 pg/mL, CSF pTau181 remained in the normal range in 14.3% (5/35) or 20.0% (7/35), respectively. CSF tTau levels were highly correlated with pTau181 (**Figure 2B**).

**Figure 2.**
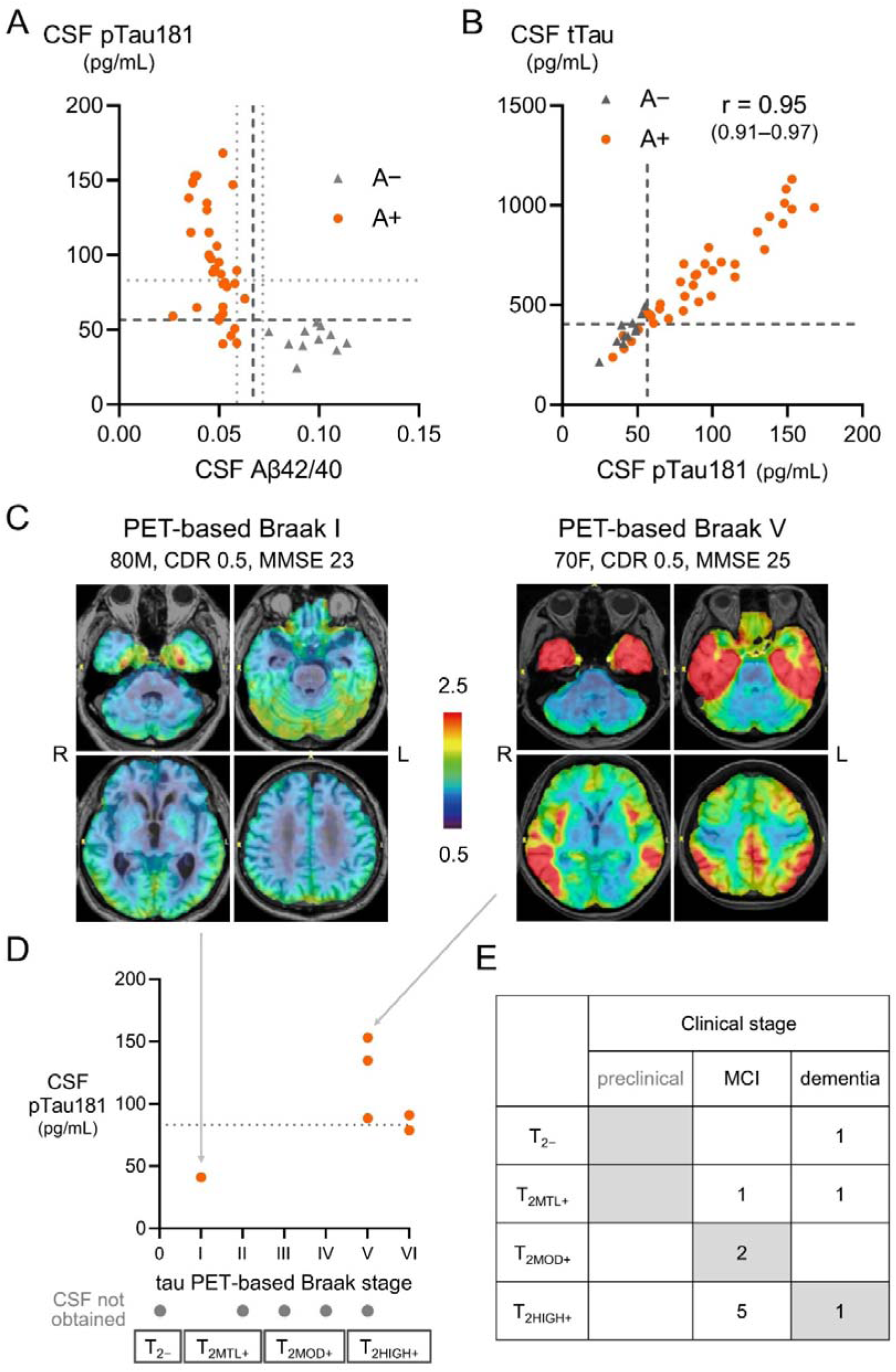
CSF biomarker results and tau PET using ^18^F-MK6240 PET A) All patients with CSF Aβ42/40 ratio above the approved cutoff of 0.067 (A−) showed pTau181 value below the cutoff (56.5 pg/mL) (both in dashed lines). Of the 35 patients with CSF Aβ42/40 below the cutoff, three (8.6%) had values close to the cutoff in the likely-positive range by the US FDA, associated with some uncertainties (0.059–0.072) (in dotted lines). These three patients had pTau181 values of 41.0 (< 56.5), 70.8 (56.5–83.1 [in dotted line]), and 89.6 (> 83.1) pg/mL, respectively. Three patients with clearly decreased Aβ42/40 ratio (< 0.059) had pTau181 values in the normal range (< 56.5 pg/mL). The remaining patients had abnormal pTau181 values above (n = 19) or below (n = 13) 83.1 pg/mL. (B) CSF tTau is highly correlated with p-tau181 (r = 0.95). (C) Representative tau PET images using ^18^F-MK6240 PET. Color scales represent the standardized uptake value ratio, with the cerebellum as the reference region (= 1). (D) Tau PET-based Braak stage ranges from 0 (normal) to VI (max). While one with PET-based Braak stage I had low CSF pTau181, patients with stage V–VI tended to have higher CSF pTau181 values (41.0 vs 109.1 ±32.7 pg/mL, p = 0.13). (E) The association between biological stages based on tau PET and clinical stages is presented. A−, amyloid negative; A+, amyloid positive; Aβ, amyloid-beta; pTau181, tau phosphorylated at threonine 181; tTau, total tau; T_2−_, T2 biomarker negative (no uptake on tau PET); T_2MTL+_, tau PET uptake restricted to medial temporal areas; T_2MOD+_, moderate tau PET uptake in the neocortex; T_2HIGH+_, high tau PET uptake in the neocortex

Tau PET (T2) using the AD tau-specific tracer ^18^F-MK6240 was performed in a subset of patients (n = 11). Representative images have been previously published [25] and are presented in **Figure 2C**. The Tau PET-based Braak stages ranged from zero (none) to VI (max) (**Figure 2D**). Both tau PET and CSF pTau181 results were available for six patients; despite the small sample size, CSF pTau181 tended to be higher in patients with tau PET-based Braak stage [–[ than those with stage [ (**Figure 2D)**. The association between biological stages based on tau-PET and clinical stages varied widely (**Figure 2E**). Five patients with biological > clinical stage tended to be younger in age (70.8 ±8.5 vs. 76.7 ±9.1 years).

### 3.3 Neurodegeneration (N), Inflammation/astrocytic activation (I), and Vascular (V) biomarkers

Plasma NfL (N) ranged from 10.0 to 103.3 pg/mL and did not differ significantly between the A+ and the A− groups (**Figure 3A**). Plasma GFAP (I) ranged from 121.9 to 652.5 pg/mL and was significantly higher than the A− group (**Figure 3B**). The patients were grouped into four quartiles (Q1, Q2, Q3, and Q4) for each biomarker. White matter hyperintensities (V) scored by the Fazekas score ranged from 0–2 but included some with 3 (max) for both DWM and PV (**Figure 3C**). The associations among plasma NfL, GFAP, Fazekas DWM, and Fazekas PV were highly heterogeneous (**Figure 3D**).

**Figure 3.**
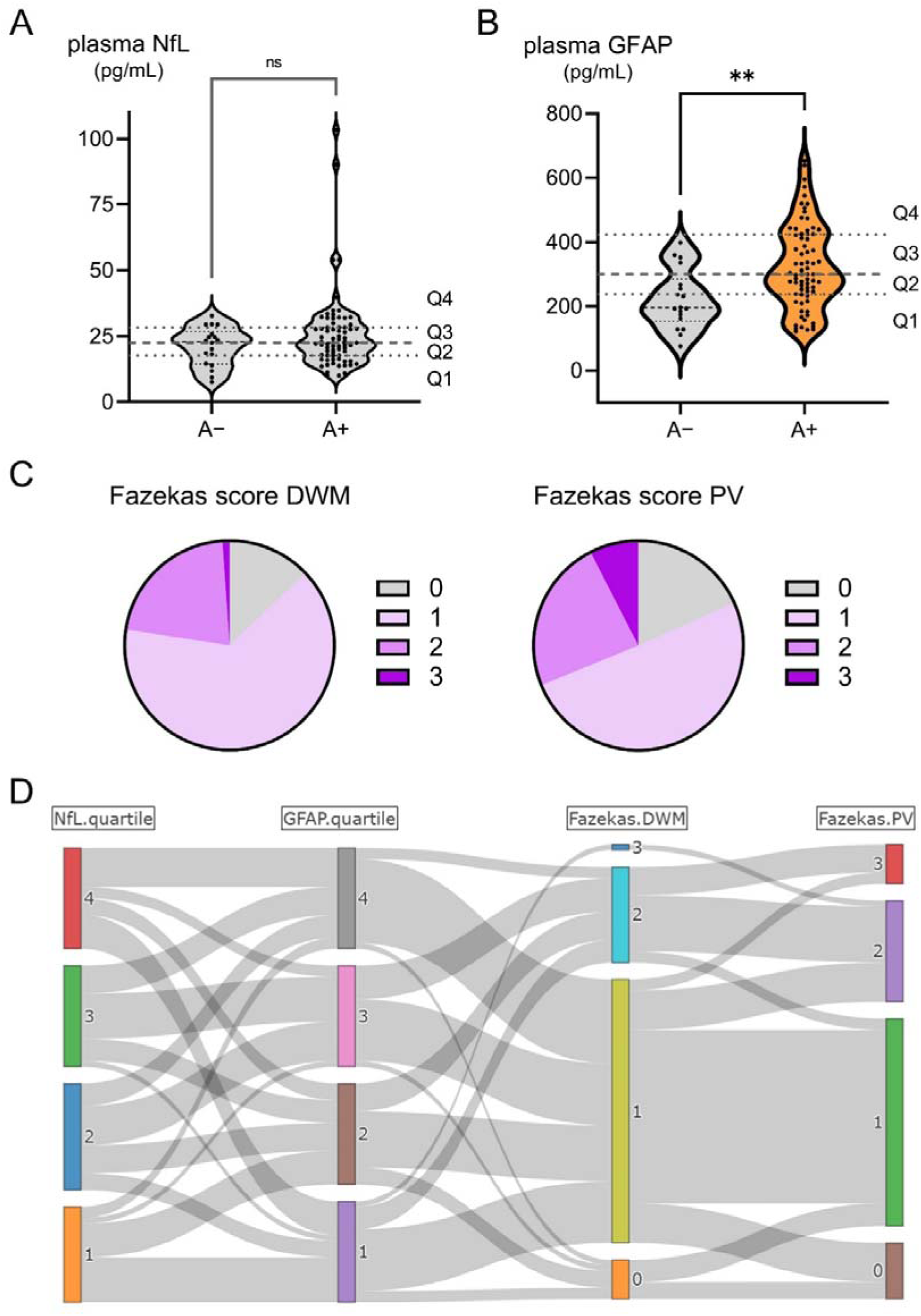
Plasma NfL, plasma GFAP, and white matter hyperintensity by Fazekas score (A, B) Plasma concentrations of NfL and GFAP were compared between A− and A+. While NfL showed no significant difference, GFAP was significantly higher in A+. Plasma NfL and GFAP showed wide distribution from 10.0 to 103.3 pg/mL and from 121.9 to 652.5 pg/mL, respectively. In the A+ group, values were grouped into four quartiles (Q1, Q2, Q3, Q4) for each biomarker. (C) White matter hyperintensities (V) scored by the Fazekas score mainly ranged from 0–2 but included some with a maximum of 3, both for DWM and PV. (D) The association between plasma NfL, plasma GFAP, Fazekas DWM, and Fazekas PV was highly heterogeneous A−, amyloid negative; A+, amyloid positive; DWM, deep white matter; GFAP, glial fibrillary acidic protein; NfL, neurofilament light chain; PV, periventricular

### 3.4 α-syn (S) biomarker

α-syn SAA (S) was evaluated in 24 patients with available CSF samples. Six (25%) patients showed clearly positive results, and two (8.3%) showed low positive results. All patients with SAA positivity were screened for signs of Lewy body disease (LBD). Only one patient showed signs of parkinsonism at evaluation, and the remaining patients showed no signs of LBD, including fluctuating cognition, visual hallucinations, REM sleep behavior disorder episodes, parkinsonism, hyposmia, lightheadedness on standing, or constipation.

### 3.5 Association between A biomarker characteristics and other TNIVS biomarkers

Although the sample size was small, we conducted an exploratory analysis to compare TNIVS biomarkers between patients grouped according to A biomarker characteristics (**Table 2**). Some associations were indicated, including higher Fazekas scores in the amyloid PET regional-positive group and higher plasma NfL in the CSF Aβ42/40 0.059–0.067 group.

## 4 Discussion

We identified substantial heterogeneity in ATNIVS biomarker results in real-world patients receiving lecanemab.

### 4.1 Heterogeneity in Core 1 (A, T_1_)

Amyloid PET burden measured by the centiloid scale was lower than that reported in the global or Asian subpopulations of the phase 3 trial (**Table 1**) [2, 27]. This discrepancy may reflect differences in PET acquisition, visual interpretation, quantification methods, and racial/ethnic differences. However, amyloid PET images were acquired using ^18^F-flutemetamol through a standard protocol, visually interpreted according to approved criteria, and calculated centiloid scales using the VIZCalc software, which is consistent with other methods [38]. Although our study included patients with low centiloid values close to 0, which could have lowered the mean value, the phase 3 trial also included those with low centiloid values, the lowest being −16.6 [2]. There are uncertainties about whether there are racial differences in the amyloid PET burden at similar clinical stages. The mean amyloid PET centiloid in the phase 3 trial of donanemab was numerically lower in Japanese (80.8–89.9 vs. 100.9–103.5 centiloid) [3, 39], although this difference was not observed in the phase 3 trial of lecanemab.

While amyloid PET (A) was visually positive in all tested patients who started lecanemab, the distribution varied as previously reported in research cohorts. Although the majority showed Aβ deposition in all five brain regions for the approved flutemetamol visual read, 21% had only regional positivity (A+_region_). A previous study in a research cohort using similar methods reported the prevalence of A+_region_ to be 24% in end-of-life patients (including asymptomatic patients) and 13% in patients with A+ amnestic MCI [40]. As these percentages were similar, there were some differences in their distribution patterns. While a previous study reported that the posterior cingulate cortex (PCC)/precuneus was most frequently positive in the A+_region_ group, the frontal lobes were most frequently positive in this study, followed by the temporal lobes and PCC/precuneus. Another flutemetamol study using a different analytical method reported that the frontal lobe was the most frequently affected brain region [41]. In this study, patients in the A+_region_ group were older and tended to have milder clinical stages. Patients with the A+_region_ may have other clinical characteristics that are important for understanding the heterogeneity in AD, and further studies are warranted.

Although CSF Aβ42/40 ratios (A) were below the approved cutoff in all tested patients who initiated lecanemab, some values were close to the cutoff in the likely-positive range warned by the US FDA. Recent studies show that CSF pTau181 increases in the presence of brain Aβ pathology, and two of the three patients with CSF Aβ42/40 ratio within this range showed increased pTau181, suggesting AD, whereas one showed a normal pTau181 value. As observed in those with clearly decreased CSF Aβ42/40 ratio, the meaning of normal CSF pTau181 remains to be determined. Our previous autopsy validation study included patients with normal CSF pTau181 levels and advanced AD pathology at autopsy [17]. Conversely, in the present study, CSF pTau181 above the cutoff was specific to those with an abnormal Aβ42/40 ratio. Although caution is needed in some rare diseases [18], our results may suggest that pTau181 increase is relatively specific for AD, at least in real-world patients evaluated for anti-Aβ antibodies.

### 4.2 Heterogeneity in Core 2 (T_2_)

Recently revised criteria divide tau biomarkers into T1 and T2 stages [6]. Tau PET is the most reliable T2 biomarker and, although not required for AD diagnosis, it is important for understanding the biological stage of AD [6]. Four biological stages have been proposed based on the tau PET distribution and uptake [6]. ^18^F-MK6240 is a second-generation tau PET tracer with high sensitivity and specificity for AD tau pathology and minimal off-target binding to the choroid plexus [42], enabling better evaluation of the medial temporal cortex [43]. Tau PET-based Braak stage using this tracer was proposed and has been used in multiple studies [21–23]. PET-based Braak stage 0 corresponds to no uptake, and stages I–II correspond to uptake restricted to the medial temporal areas, which may not be directly associated with MCI or dementia stages [6]. Although the revised criteria proposed using the uptake magnitude in the neocortex to separate the latter two stages (moderate vs. high), some of the earliest reports used a topography similar to the PET-based Braak stage to approximate the discrimination of these two stages [44].

In this real-world cohort of patients with MCI or mild AD dementia treated with lecanemab, tau PET-based Braak stage ranged from normal (0) to fully abnormal (VI). Three patients (27%) with PET-based Braak stages 0–II showed more advanced clinical stages than those suspected from the biological stages, and may suggest co-pathologies [6]. This proportion is consistent with recent reports that used different tau PET methods [44–46]. Five patients with PET-based Braak stages V–VI had MCI. Some of these patients may have a biological stage that is more advanced than that suspected based on the clinical stage, and may suggest resilience/reserve [6]. Continuing research in real-world patients receiving anti-Aβ antibodies should be important to understand the heterogeneity in tau pathology.

CSF pTau181 is known to increase with Aβ pathology and is also associated with AD tau pathology/PET tracer uptake in individuals with A+ [17, 23]. Previous studies suggested that CSF pTau181 is further increased at Braak stages III–IV, but does not continue to increase in later Braak stages (V–VI) [23]. Although only a limited number of patients had both CSF pTau181 and tau PET results, patients with PET-based Braak stages V–VI in this study also tended to have a higher CSF pTau181, mostly above our previously estimated cut-off [14, 17]. While waiting for approval of better biomarkers for AD tau pathology/PET tracer uptake, measuring CSF pTau181 in addition to the Aβ42/40 ratio, may have some additional benefits.

### 4.3 Heterogeneity in NIV

Neurodegeneration (N) and inflammation/astrocytic activation (I) biomarkers are nonspecific to AD, but are important in AD pathogenesis [6]. CSF/plasma NfL and GFAP are the most common fluid biomarkers for assessing N and I. Cerebrovascular disease is a common and important co-pathology of AD. Although there is no consensus on which indices should be used as vascular (V) biomarkers, white matter hyperintensities (WMH) on MRI are often evaluated, and may partly reflect microvascular injuries [6]. The Fazekas score is widely used for scoring the severity of WMH. High intensities in the PV and DWM are scored separately and may differ in etiology and functional correlates [47]. In this study, we identified substantial heterogeneity in N, I, and V biomarkers and their combinations. Based on the results of previous studies [6, 48], these biomarkers may become important for understanding the heterogeneity in disease progression speed and other follow-up variables in future studies using real-world data.

### 4.4 Heterogeneity in S (α-syn)

Lewy bodies, composed of α-syn, are a common co-pathology in patients with AD [49, 50]. Now that highly-specific α-syn SAAs are available, studies have started to test in patients with AD [50] or in a wider population, including AD [51, 52]. Positivity has been reported to be 10–30% in patients with AD [19, 50, 53–55] and may be associated with distinct features [52, 56]. However, the positivity rate of lecanemab in real-world patients remains unknown.

In this study, 25–33% were α-syn SAA positive using a previously validated method [19]. The positivity rate was slightly higher than that in some studies on AD [19, 54, 55], but was within the reported range. This discrepancy could be because of many reasons, including differences in the study population, assay, or cutoff values. However, the fact that the same method yielded lower positivity (10%) in our previous study [19], differences in the study population, such as higher age (65.2 ±11.6 vs. 74.2 ±8.1 years old), may have contributed. Importantly, only 12.5% (1/8) of α-syn SAA-positive patients showed clinical signs of LBD at evaluation. Our results further support the importance of evaluating α-syn SAA even in patients without obvious symptoms of LBD.

### 4.5 Association between A and other biomarkers

Despite the small sample size, there were some indications from the exploratory analyses comparing A+_region_ vs A+_wide_ by amyloid PET and between likely- (0.059–0.067) and clearly- (< 0.059) positive by CSF Aβ42/40 ratio. The Fazekas scores (both DWM and PV) were higher in the A+_region_. This difference may be explained by vascular factors contributing more often to cognitive symptoms in the A+_region_ group. Fazekas scores also tended to be higher in the CSF Aβ42/40 0.059–0.067 group. Plasma NfL was higher in the CSF Aβ42/40 0.059–0.067 group. Although the reasons remain unknown, the results suggest that more severe neurodegeneration caused by AD or co-pathologies is associated with higher plasma NfL levels. Although based on very few cases, the tau-PET-based Braak stage tended to be lower in these groups. Patients with A biomarkers closer to the threshold may tend to have a lower tau burden and more co-pathologies that contribute to cognitive symptoms. However, caution is necessary because there have been reports of low amyloid PET and high neocortical tau PET [20, 57].

### 4.6 Limitations and future perspectives

This study has some limitations. First, this was a retrospective study using data obtained from a clinical setting, and there were missing results for several biomarkers (particularly T1, T2, and S). Since most patients donated their blood samples to our biobank, future studies using blood-based biomarkers, including T1 (pTau217), T2 (MTBR-tau243) [58], and S (α-syn SAA) [59] would be important. Second, this was a single-center study conducted in Japan, and each institution and country should have different populations and situations. Larger nationwide and worldwide studies are important to understand the full picture of the real world. Third, because data collection was ongoing, we summarized baseline data and did not include follow-up data in this report. We plan to report the association between ATNIVS biomarker heterogeneity and follow-up data (clinical outcomes and adverse events) as soon as we obtain sufficient data.

## 5 Conclusion

We identified substantial heterogeneity in ATNIVS biomarkers among real-world patients receiving lecanemab. These results are important for understanding heterogeneity in future analyses using real-world data.

## Data Availability Statement

The data supporting the findings of this study are available from the corresponding author upon reasonable request.

## Acknowledgments

The authors thank all patients and their family for their participation in our various studies. We also thank members of the Memory Clinic; Department of Neurology/Psychiatry/Nursing/Clinical Psychology/Radiology; Dementia Support Center/ Regional Medical Coordination Office; Healthy Aging Innovation Center (HAIC), TMIG biobank, and the Integrated Research Initiative for Living Well with Dementia (IRIDE) for their assistance.

